# Revisiting the use and effectiveness of patient-held records in rural Malawi

**DOI:** 10.1101/2025.03.01.25322650

**Authors:** Amelia Taylor, Paul Kazembe

## Abstract

**Background:** Health Passports (HPs) are paper-based, patient-held records used in Malawi to document key details about the health condition of a patient and the care provided during medical visits.

**Aim:** This paper assessed their use and effectiveness within the health data ecosystem, and their impact on patient care.

**Setting:** The study setting was facilities in the Zomba District, Malawi.

**Methods:** We undertook a descriptive exploratory qualitative study to determine the practices for data recording used by health care professionals and the importance placed on HPs by patients and professionals. Pages from completed HPs were analysed to extract practices for recording case presentation, diagnosis, medication.

**Results:** Despite their significance, there was a deterioration in their use. HPs are mainly used in government clinics and for the poorer segment of the population which does not have access to private healthcare and insurance. The rural population is also affected by deteriorating literacy and health infrastructure. Inadequate practices in recording patient notes negatively affected their effectiveness as source of information for patients and health professionals.

**Conclusion:** There is a need for health policies and systems to recognise the importance of HPs and direct efforts to support their effective use. Handwritten HPs have great potential if the clarity and consistency of the written communications is improved. Guidelines for record keeping are needed to help improve the effectiveness of HPs so that they work with existing laws for the benefit of patients and professionals.

**Contribution:** This study contributes to an under-researched area for determining the effectiveness of patient-held records in LMICs.

## Background

The use of HPs in all health facilities in Malawi is required by the Malawi Health Information Systems Policy: patients must present their HP at each visit and healthcare providers must use it to record key details about the patient presentation, diagnosis, care plan, treatment, and follow-up dates (1). The use of HPs enables a decentralised and portable health information system, as they allow patient records to be accessible to any healthcare professional, regardless of the hospital or facility the patient visits. The health information system landscape in Malawi has changed considerably in the last 25 years since HPs have been introduced in early 2000s. The number of records that health professionals need to complete for each patient has increased and many rural facilities do not have adequate resources to maintain facility based records (2).

Notably, recent digital health initiatives tend to focus on introducing electronic systems whose role is to aggregate data collected through manual means at rural facilities in districts and whose implementation is mainly possible in urban, private or larger government facilities (3,4). In Malawi and many other African countries, paper-based records continue to be used alongside electronic records (5). A hierarchical system of data collection, data transcription facilitates the recording and transmission of data from rural clinics, district facilities and then to the national level (4). In rural clinics most data are collected and recorded on paper. HPs are also used by patients referred to larger government hospitals in urban centres.

Paper-based records are frequently used to check the validity of data captured in electronic systems (7). If a decline exists in the record-keeping practices in Malawian healthcare facilities, this will not only impact the quality of patient care but will also hinder healthcare professionals from transitioning to and upskilling in electronic record systems. Poor record management can lead to inaccurate diagnoses, delays in treatment, inefficiencies, and data gaps that affect decision-making. Weak paper-based records also make digitisation more challenging. As early as 2014, it was noted that a lack of HPs, due to loss or damage, or inefficient use by health professionals constituted a challenge in moving to digital EHRs due to loss of historical patient data (8). HPs together with other paper based registers provide valuable patient data, for many people in Malawi they are the only available medical record, hence the information they provide is crucial for the digitalisation of healthcare in Malawi.

The basis for our investigation was a study we conducted into language barriers in communication between health practitioners and patients in facilities in Malawi (9). We observed that Health Passports (HPs) played a significant role during health visits but that it was not clear how the data recorded in them was utilised by healthcare professionals and patients. We found that many patients did not understand or were confused by what was written in their notes. Part of the problem stemmed from the use of English and low patient literacy; however, these factors alone did not explain why healthcare practitioners failed to utilise patient recorded history in HPs when making diagnoses. These findings pointed to the need to revisit the utilisation and effectiveness use of HPs, and the importance placed on them by patients and health professionals.

This study engages with healthcare professionals in rural clinics and with literature to assess the effectiveness, usage, and value of HPs for practitioners, patients, and the healthcare system. It also aims to identify key barriers that hinder their effective use.

## A brief history of HPs in Africa

The earliest type of health passports introduced in Africa recorded and tracked maternal and child health (10). These health passports varied in format and complexity, ranging from simple antenatal notes or vaccination cards to more comprehensive health booklets. They have been known under many names such as “maternal and child health booklets” in Kenya (11), “home-based maternal records” in Zimbabwe (12) or “women-held notes” or “ante-natal cards” in Zambia (13), “child health cards” in Uganda (14), “road to health cards” in South Africa (15). Progressively booklets for general health were introduced. We found several studies that mention the use of HPs in various countries in Africa although these studies do not provide detailed insights into formats, guidelines or effectiveness of HPs over time.

In developed countries, patient-held records are typically copies of the records kept about the patient at facilities, their primary purpose being to inform patients and encourage their participation in their care plan. Suitability for different age groups in terms of the information captured in the notes, a lack of agreement between patients and health professionals regarding their function and confusion over ownership of the records negatively influenced their adoption in developed countries (16). In Africa, studies on HP tend to emphasise their importance and on positive impact on patient care (17). Placing health records in the custody of patients seemed to be a good solution to resource limitations faced by health systems [10]. In the absence of integrated systems capable of sharing patient data between facilities, HPs were envisaged to be a mobile health record presented and used at any visit and any facility by the patient.

## Methods

### Design

We use a descriptive exploratory qualitative design informed by a study we conducted previously into language barriers in health facilities in Zomba District (9). The lines of inquiry used during the Focus Group Discussion (FGD) are listed in *Table 1*. Participants for FGD were selected from the first study on the basis that they were knowledgeable about the subject matter and held leadership positions in their respective facilities. The research was conducted between September 2024 and January 2025. We used document analysis to assess the quality of the notes recorded in HPs.

**Table 1.** Study questions and dimensions of inquiry.

| Dimension | Questions considered |
| --- | --- |
| **History** | When were HPs introduced in Malawi? |
| **Provider Use** | How do healthcare professionals use the information in the HP?<br>Have you done any training on how to record notes, or how to make entries in the HPs?<br>Are there symbols that are commonly used? |
| **Importance** | What value do you think healthcare professionals place on HPs?<br>What value do you think patients place on HPs? |
| **Effectiveness** | Do patients ask for clarifications about what is written on their HPs?<br>Do you discuss with patients what is written on their HPs during consultation?<br>How could HPs become more useful to healthcare professionals and patients? |
| **Impact of literacy** | What do you think is the impact of low literacy on the quality of information exchanged during consultations?<br>In what way do you think low literacy affect the ability of patients to express their symptoms and conditions?<br>In what way do you think literacy could or does affect the ability of patients to express to understand their diagnosis and treatment? |
| **Language Use** | Do you think there is a case for writing symptoms and diagnoses in Chichewa or the language of the patient in the HP? |
| **Issues of Privacy** | To what extent do you think a lack of privacy affects good communication?<br>In what way do you think a lack of privacy affects the quality of healthcare?<br>What kind of privacy situations occur?<br>What can be done? |
| **Link to eHealth Systems** | How are HPs used in connection with registers, tally sheets at facilities?<br>How are HPs used in relation to other electronic records?<br>How does their use impact training and readiness for full digital health systems? |
| **Ownership** | Who owns the patients' health records? |

The study setting was rural facilities in Zomba district. Ten healthcare professionals participated in the FGD: 3 were women and 8 were men. They represented 9 different clinics from the district, all being situated in rural areas, except a clinic in Zomba town who served both urban and rural patients. One, H. Parker, was a private clinic; the rest were government run. Among participants were clinic heads, head clinicians, nurses, data clerks, and technicians with varying number of years of experience in the health sector.

The data we analysed in this study comprised of:

- Sample images from HPs that were obtained from patients who attended clinics in Zomba District in the first study. These were used to determine the practices of health professionals in recording patient notes and the quality of the coding used. These are presented in the Supplementary material for the paper.
- Transcriptions of audio recordings and notes taken during the FGD provided key research themes. We analysed the themes along the lines of inquiry in *Table 1*.
- Policy and guideline documents were consulted to establish the existence of guidelines and procedures for recording patient notes.

The use of HPs is clearly stated in the National Health Information System Policy (NHIS) (1). Table 2 contains the paragraphs from the NHIS mentioning the use of HPs. Health practitioners are required to update the HP during each visit with essential medical information from patient records maintained and kept at facilities. The policy also specifies the type of information that should be recorded in HPs: proposed diagnosis, plan of care, treatment given and follow-up date. The NHIS policy has not been updated since 2015 and does not cover practical considerations on how to record health notes.

**Table 2.**
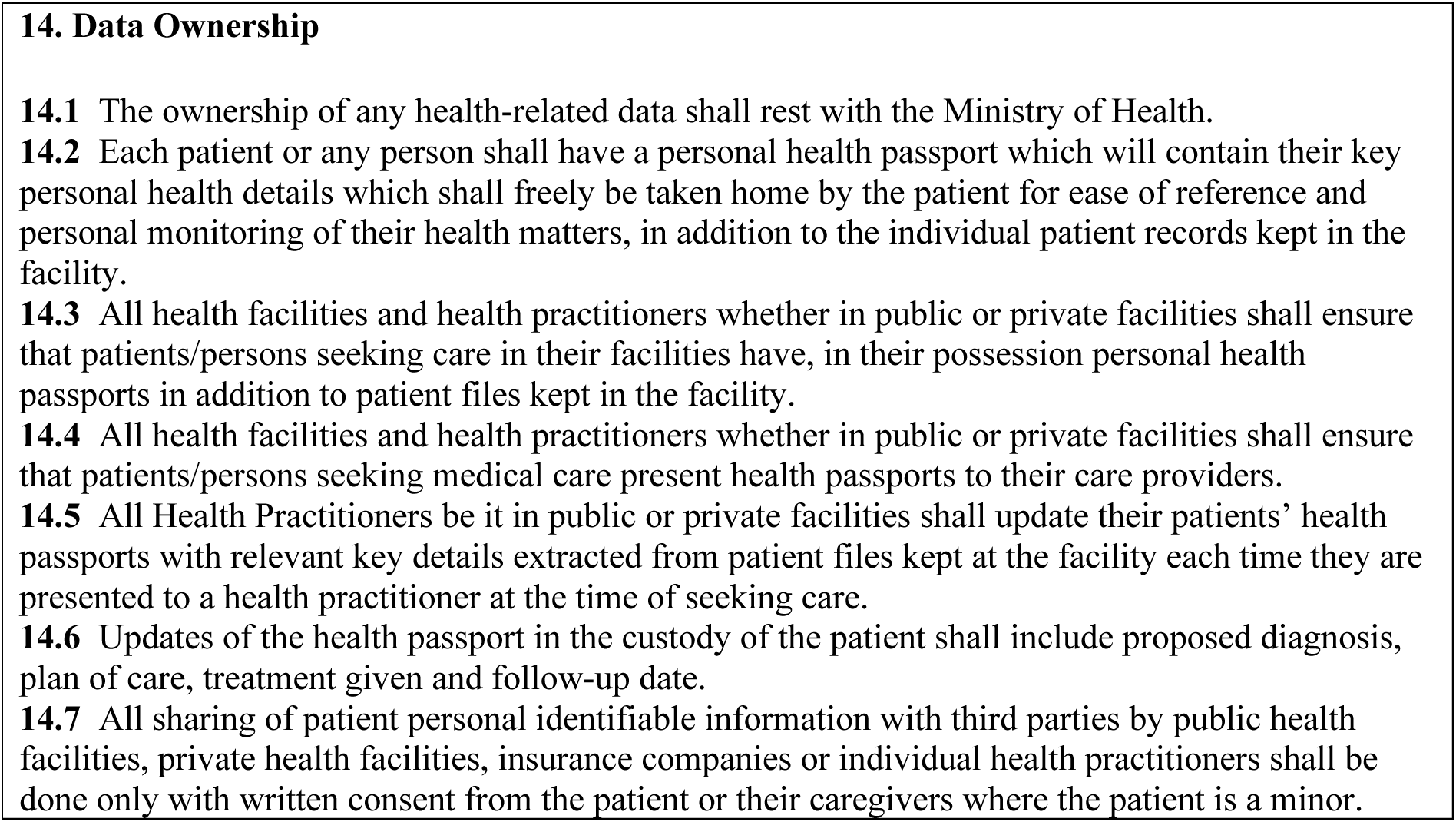
Data Ownership and use of health passports as per the Malawi National Health Information System Policy 2015.

|  |
| --- |
| **14. Data Ownership**<br>**14.1** The ownership of any health-related data shall rest with the Ministry of Health.<br>**14.2** Each patient or any person shall have a personal health passport which will contain their key personal health details which shall freely be taken home by the patient for ease of reference and personal monitoring of their health matters, in addition to the individual patient records kept in the facility.<br>**14.3** All health facilities and health practitioners whether in public or private facilities shall ensure that patients/persons seeking care in their facilities have, in their possession personal health passports in addition to patient files kept in the facility.<br>**14.4** All health facilities and health practitioners whether in public or private facilities shall ensure that patients/persons seeking medical care present health passports to their care providers.<br>**14.5** All Health Practitioners be it in public or private facilities shall update their patients' health passports with relevant key details extracted from patient files kept at the facility each time they are presented to a health practitioner at the time of seeking care.<br>**14.6** Updates of the health passport in the custody of the patient shall include proposed diagnosis, plan of care, treatment given and follow-up date.<br>**14.7** All sharing of patient personal identifiable information with third parties by public health facilities, private health facilities, insurance companies or individual health practitioners shall be done only with written consent from the patient or their caregivers where the patient is a minor. |

We found that the following procedure for data collection was used in the facilities we visited. Data clerks had a dedicated desk space and collected data from patients finished after consultation. They transcribed data from HPs into OPD (Out-patients department) and other registers according to the services that the client received. The OPD register captured the name, age, sex, symptoms and treatment.

Therefore, because HPs are a source of health data for other systems, the impact of poor records in HPs could be significant. Generally, data from facility registers is compiled, aggregated and reported monthly using both programme-specific reports (e.g. maternity, ART, etc.) and composite reports. Data collection and aggregation is the responsibility of Health Management Information Systems officers who oversee the work of the data clerks. Malawi uses electronic systems for aggregating routine facility and/or community service data (4).

### Ethical clearance

The study was approved by the Research Committee of the Zomba DHO (ZA/DC/ADMIN/15/02/2024). All participants were sent an invitation clearly explaining the purpose of the FGD and gave their consent in writing.

## Results

### Primary outcomes

#### The Format of HPs has remained largely unchanged since their introduction

We found that the official HPs format and colour scheme has remained largely unchanged over time since their introduction by the Fourth National Health Plan 1999-2004 (18). But currently the format and quality are not controlled. Patients can buy HPs from vendors who are selling them outsides of clinics. Participants in the FGD recalled a time when health passports were issued only by recognised organisations and had a recommended number of pages and quality. Presently many HPs have very few pages limiting their use during multiple visits. HP sell at K800, and their cost has become a burden and a deterrent for many poor patients.

#### Insufficient patient medical information was recorded

We found that the general history section was blank for most patients. X-rays and other test results are not routinely provided to patients or attached to the HP. The cover page of the health passport includes the patient’s name, date of birth, village, traditional authority (T/A), and district, which can be filled in by either the patient or the healthcare professional (Figure 1 and Figure 2). A section on general history divided into three parts found on the first pages contains the medical history of the client. Part A is for the psychosocial history, which includes occupation, marital status, alcohol and tobacco use, religion, known allergies, and blood group. Part B contains family history, including known allergies, mental health conditions, asthma, and other relevant conditions. Part C is dedicated to past medical and surgical history.

**Figure 1.**
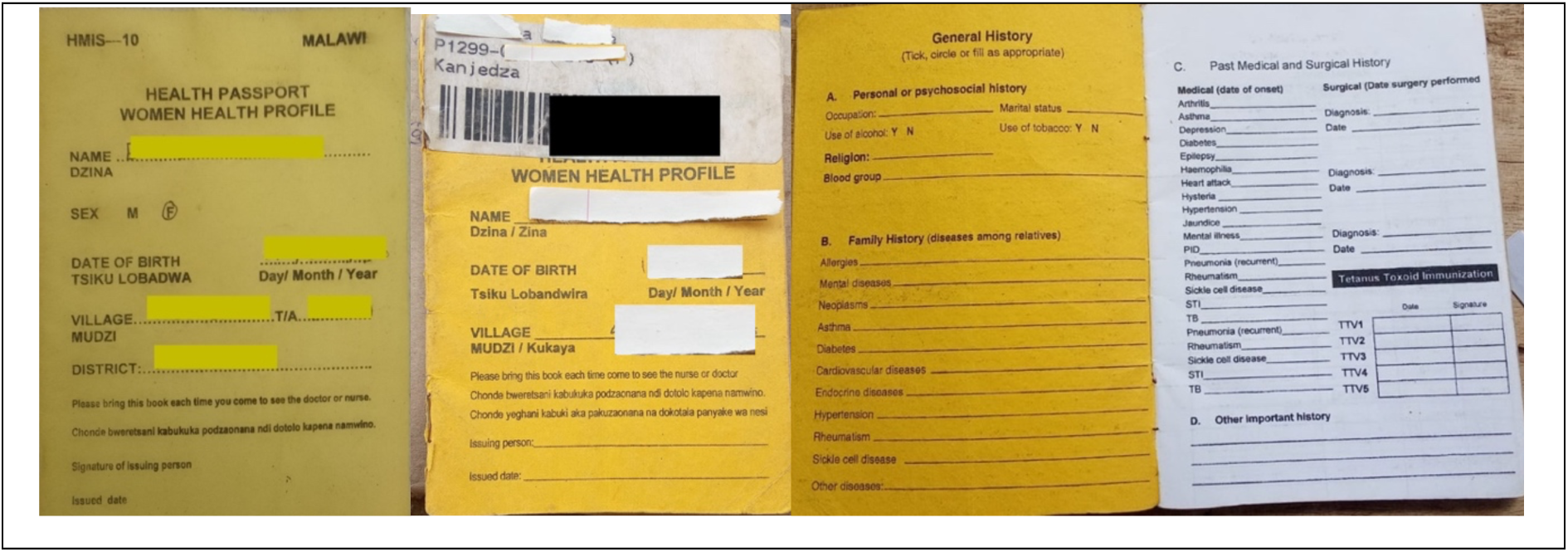
Samples HPs for women (yellow cover), cover and inside cover and front sheet. The HP contains standardised sections for immunisation, family planning, previous deliveries. A girl is given a women’s health booklet when she reaches puberty. For small girls the HP is pink and is issued at birth, and contain standardised sections for vitamin A, immunisation and growth monitoring.

**Figure 2.**
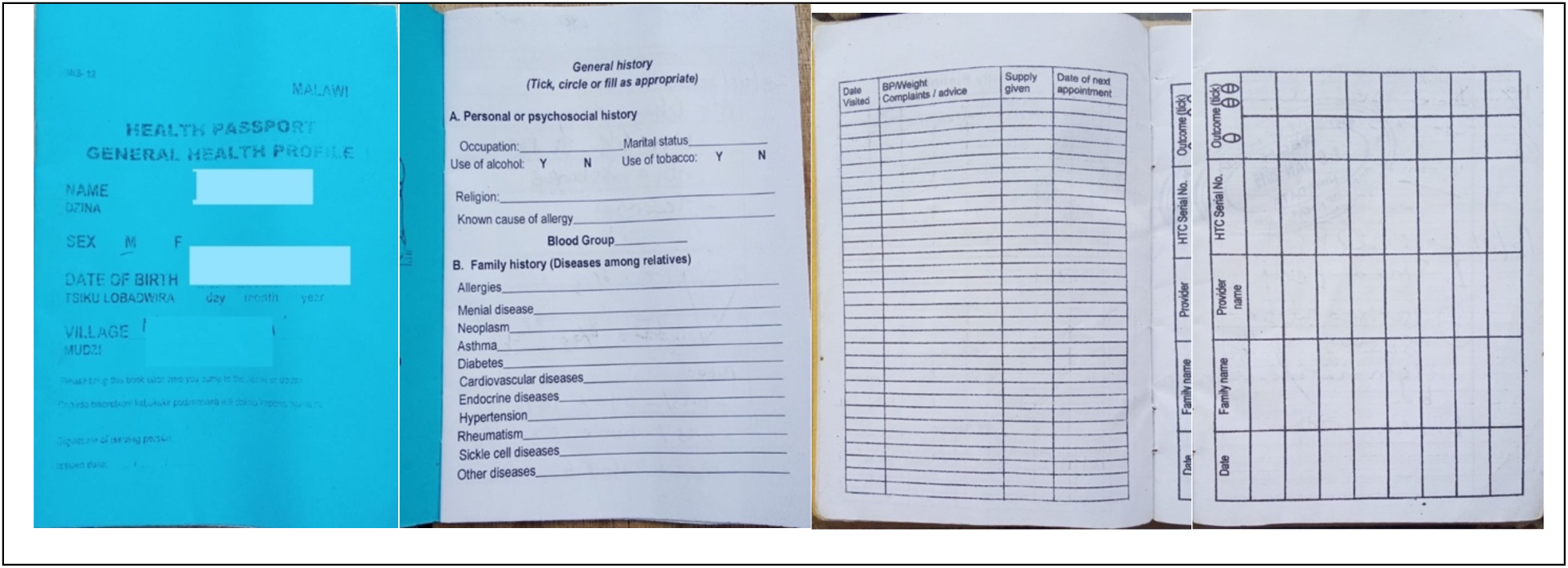
Samples of new, probably unofficial (above) and original HPs for men (blue, below). S general health booklet for male adults is usually annexed to the child health booklet for continuous recording of assessment and care.

### Completeness of records and provider use

Participants said that they used the HP regularly, but there were exceptions. In private clinics and those that served clients on private health insurance schemes, HPs were not used. Instead, patients received printouts or handwritten notes with test results and medication prescriptions, often tied to billing. In these cases, facility records were linked to insurance schemes, which had their own forms for registering patient visits and documenting care. This omission was seen as negligent, as it contributed to gaps in medical history and undermined the HP’s role as a comprehensive health record. We found that many of the standardised pages, for example, history of medical conditions or procedures and the table for listing past visits were rarely used (Figure 3 and Figure 4).

**Figure 3.**
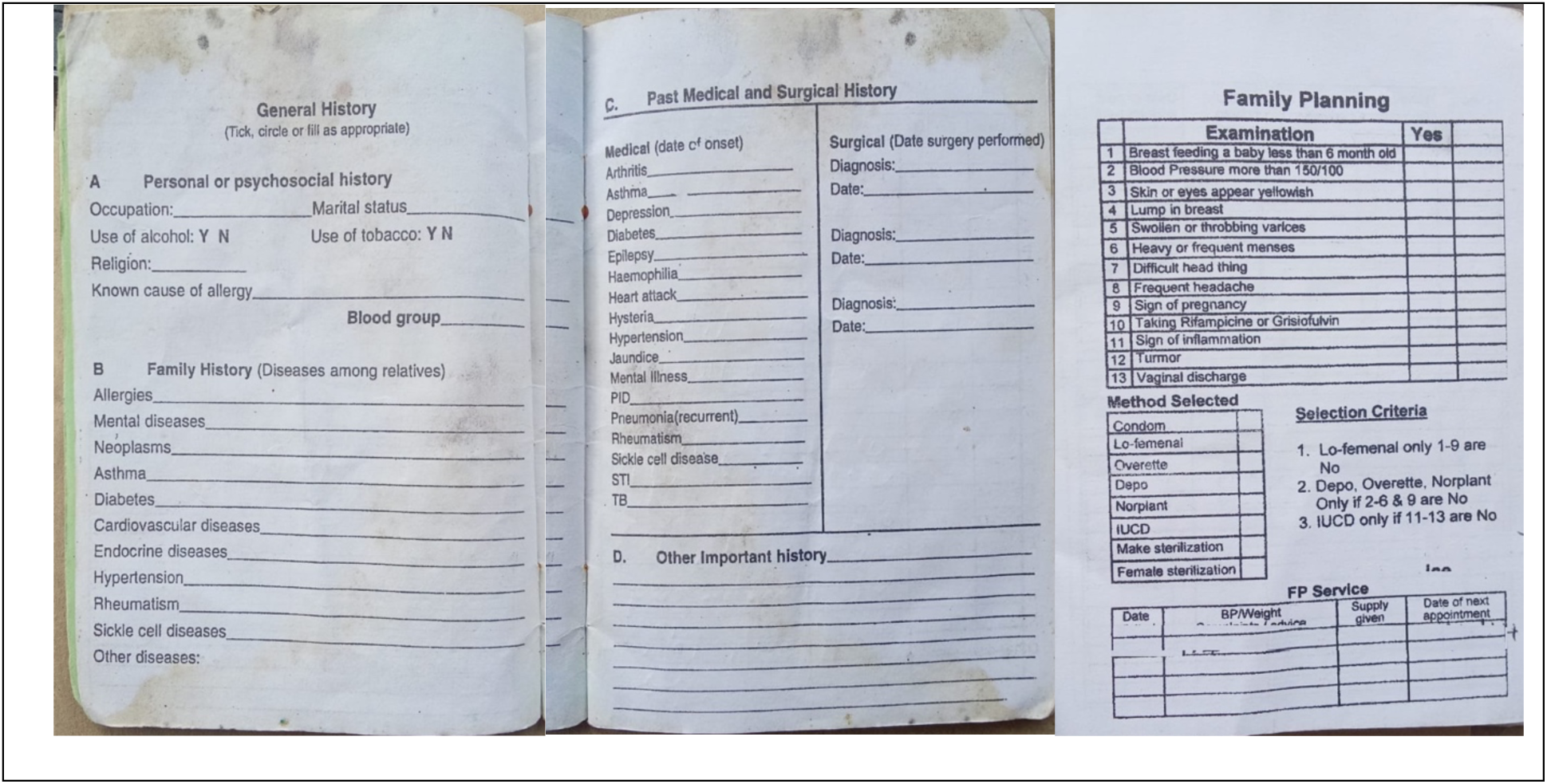
Examples of standardised pages in general care HPs

**Figure 4.**
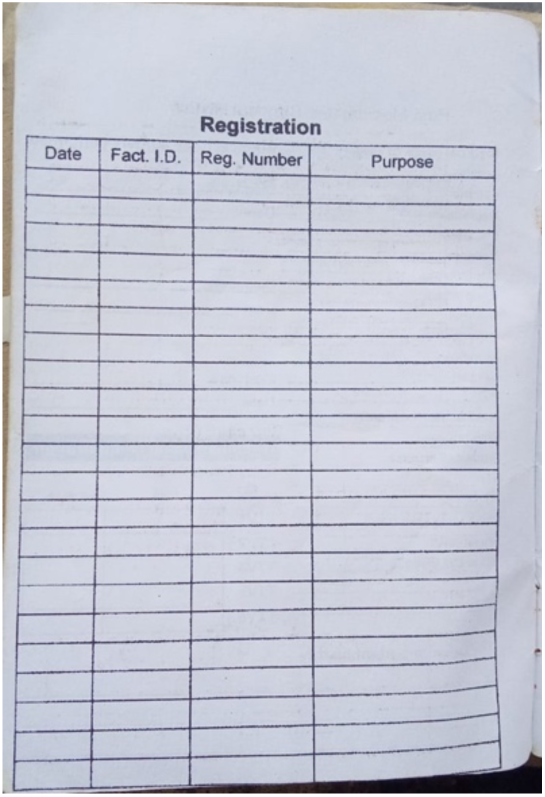
Page to record visits to any health facility. The registration number can be generated by a facility, or it can represent an insurance number. Fact I.D. may refer to a specific treatment, e.g. vaccine.

Participants agreed that patients generally did not value their HPs and saw them primarily as a requirement for accessing healthcare. Women and elderly patients were more likely to bring their HPs on a more regular basis. Cases of damaged, lost, or misplaced HPs are common. Patients had limited awareness of the importance of maintaining a single, reliable health record. Many professionals admitted feeling guilty about not educating patients on the importance of the HP. Participants identified several common problems and situations:

- some patients did not have a health passport, as majority of local villagers did not afford to buy it at K800.

- some patients had several HPs. Those on Antiretroviral therapy (ART) for treating HIV infections often had a separate confidential HP for that specific condition, which along with other treatment cards were sometimes kept at ART clinics.

Notably, the ART HP was only presented to ART health officers and never to other clinicians, meaning an ART patient may have had multiple HPs.

- some patients intentionally left their HPs at home to avoid discussions about treatment or underlying conditions. This was also common among those reluctant to follow up on their prescribed care.

- some patients accepted their HP to be used by other relatives.

- private clinics did not ask patients to present their HPs during consultation because they make recordings in insurance cards or electronical records

- all entries are done in English and usually using medical terms that cannot be well understood by most patients.

#### Affordability and access issues

Concern about costs were frequently mentioned, with some patients resorting to borrowing HPs from others, for example, a man might use his wife’s HP. The NHIS does not explicitly state that care will be denied if a patient arrives at the facility without their HP, but places responsibility on health facilities to ensure that patients present their HP during visits. Some professionals refuse to see patients that do not present their HPs for consultation.

#### The relevance of HPs for arriving at a correct diagnosis

An important purpose for using HPs is to support the process of shared decision making by which practitioners and patients make informed decision on diagnostic and treatment. HPs can therefore put reliable information in patients’ hand, and this information could open discussions and information sharing between patients and practitioners during visits. We found that due to language barriers and low literacy, this purpose was hard to achieve. Patients did not understand much of what is written in their health passports. Usually, the information that is prioritised by both professionals and patients is the prescribed treatment plan.

> *“Lay patients i.e. villagers and community members including some health workers cannot understand some or all medical entries made in the health books. The illiterates remain the main victim of this because majority of them never understand what is written in their health books. Patients will only just see the medicine given such as PCM (paracetamol), Panadol, metronidazole etc, but in majority do not know how the doctor’s conclusive diagnosis was drawn from the symptoms explained. They don’t know what they are suffering from.”*

While health professionals generally view HPs as more useful to them than to their patients, they do not habitually refer to past entries in the HPs. Practitioners tended to rely primarily on the patient’s presentation during the visit. The patient’s account was considered more important than indirect explanations of symptoms provided by a guardian, as it was believed that such explanations could be prone to exaggeration.

The perception and attitude of some professionals was also regarded as a barrier to engagement with the HPs because some professionals preferred to maintain control over consultations and resisted being guided by the information in the HP. Healthcare professionals tended to prioritise communicating drug instructions and treatment schedules rather than explaining medical conditions to their patients, an approach that was observed in patients’ own focus on accessing treatment. Power dynamics also came into play; some practitioners dismiss patients who hand over their HP unopened, telling them to “bring it already opened to the right empty page (*ubweretse potsegula kale pokuti ndilembe*).” While this may be intended to save time, it also suggests that practitioners might be unintentionally signalling to patients that their medical history is not important.

#### Do patients ask for clarifications about what is written in their health passports?

Majority of patients did not ask for clarification on the entries that a doctor made in the health pass books, due to:

- poor rapport between a patient and the doctor
- the doctors might appear to do things in a hurry when serving long queues of patients
- low level of literacy among majority of the patients
- loss of confidence and low self -esteem among many patients who in majority are illiterate.

A breakdown in trust during consultation may be deduced from these findings. Some discussions about medication administration may take place with the pharmacist, who provides instructions based on the doctor’s prescription. Some patients have preferences for certain medications, often due to past experiences. This is especially common in private clinics. For example, a patient who feels symptoms of malaria may insist on being prescribed anti-malaria medication, even if the lab results are negative.

> *“Sometimes when a medication is out of stock, the patients might be given (by the pharmacist) a different drug than the one prescribed in the HP.” (P6)*

Some patients might refuse to comply with taking the dispensed drug in case the one prescribed was not available.

#### Special needs patients

Communication between healthcare providers and patients with special needs was noted as a significant challenge during consultations. Some sign language skills, such as gestures, were partially covered during medical training, but was not considered to adequately equip professionals to communicate effectively with special needs patients.

### The content of HPs

We analysed 61 unique sample pages of completed pages in several health passports. Note of these images contain patient-identifiable information. We found that various codes and abbreviations or signs were used, and that agreement was only observed in the codes used for medication.

#### How do doctors and healthcare providers make entries in HPs?

Notes in HPs were written by hand, many contained illegible handwriting, and the coding used was inconsistent. We showed several sample pages from HPs to participants during the FGD and most agreed that the legibility of the writing was inadequate.

#### Limited Space and High Workload

HPs have limited pages, a cause of brief and sometimes incomplete notes. Additionally, healthcare professionals handle large patient volumes, leaving little time to provide detailed explanations about diagnoses and conditions.

#### Use of shortcuts

Opinions on the use of shortcuts were divided. Some believed that all words should be written in full to ensure that the patient and other professionals can easily understand the information. Some referred to the use of codes in filling in registers and deduced that this should also apply to notes made in HPs. Notes were also seen as a mark of experience and were used by consultants rather than by junior staff who tended to write longer notes using full words.

> *“If you meet a consultant, they advise you to write clearly in the health passport. They write in professional language which nurses giving the medication will understand.”*

> *“This was something that was learnt in school and is acceptable by law.”*

> *“In the OPR, information is written in short form which is acceptable by government.”*

> *“The Malawi Standards for Treatments Guidelines accepts the short forms…one needs to refresh one’s memory.”*

A rare incident was recounted in which a patient insisted that his notes were written in full in his HP and requested that the health professional rewrite them to ensure no shortcuts were used.

Participants remarked that the use of long expressions and full description of diagnosis cannot be perfectly achieved due to the current design and length of the HP and time constraints. They also believed that the use of shortcuts was acceptable. One participant disagreed and was of the view that long queues or a lack of page space should not prevent health professionals from make fully understandable entries in the health passport.

> *‘And because of the long queues experienced in clinics, heathy centers and hospitals ‘we make a lot of short cuts’ (use of signs/ short medical expressions) aiming at serving many by just ensuring that the long queues are all deflated and served regardless of the quality of service offered and we usually provide a not fully articulated information in the health passbook. Though it is an important information packed book, the spaces in it are so limited ‘its brief, we can’t write everything’ only to accommodate short forms of writings in making entries.’*

Table 3 is an example of notes transcribed from a patient HP. We can note that symptoms are written mainly in full words in a section denoted by “Cl” probably standing for “Clinical”.

**Table 3.** Example of clinician notes in a health passport transcribed from the original handwriting.

|  |
| --- |
| 10/03/2023 |
| CI: Abdominal pains |
| Loose stools 4 times |
| Acute diarrhoea |
| P: DCN 200MG tds x 3/7 |
| ORS 4 sachets |

Treatment appears in a section denoted by the letter “P” (for plan) and there are codes and symbols for drugs and dosages. The date of the visit was recorded on the top left of the page.

#### Symbols used

Symbols used fell into seven categories of abbreviations: charting (13) and drug-related (11) were most common, followed by diagnosis or condition (8), and laboratory (7), pharmacy (5), vital signs (5) and anatomy (1) abbreviations. In Table 4 we counted how many different symbols in each category.

**Table 4.** Types of abbreviations used.

| Type | No. Abbreviations |
| --- | --- |
| Charting | 13 |
| Drug | 11 |
| Diagnosis / Condition | 8 |
| Laboratory | 7 |
| Pharmacy | 5 |
| Vital Signs | 5 |
| Anatomy | 1 |
| Total | **50** |

In Table 5 we list all the abbreviations found in the data alongside their most likely meaning. For example, there were 12 different symbols used for charting, e.g. Hx (History of) and C/O (Chief Complaint). Charting abbreviations referred to taking down the case history of a case but were also used to demarcate sections such as those dedicated to treatment plans and diagnosis. Not all abbreviations were used consistently: for example, some referred to treatment plan using the word Plan, others used the letter P and others used the more generally accepted “Rx”.

**Table 5.** List of abbreviations found in HPs we sampled. The starred abbreviations appear in the list of abbreviation in the MSTG.

| Notation | Meaning of abbreviation | Type |
| --- | --- | --- |
| **ABD** | Short for abdominal, e.g. abdominal pains. | Anatomy |
| **C/O** | Short for “complains of” | Charting |
| **CR** | It could stand for “Clinical Response” | Charting |
| **D** | Diagnostic (instead of Dx). E.g. D septicaemia | Charting |
| **DOA** | Date of Admission | Charting |
| **HX** | History, e.g. HX of asthma = a history of asthma | Charting |
| **O/E** | On Examination, e.g. O/E stable | Charting |
| **P** | Plan (meaning Treatment Plan) | Charting |
| **PE** | Physical Examination, E.g. hoarseness of voice | Charting |
| **PTC** | Probably a mistake, meaning PC = Presenting Complaint | Charting |
| **RC** | Possibly Related Condition | Charting |
| **RX** | Treatment | Charting |
| **X** | Diagnosis | Charting |
| **Δ** | diagnosis; change | Charting |
| **ARI\*** | Acute Respiratory Infection | Diagnosis / Condition |
| **BH** | Not clear. | Diagnosis / Condition |
| **ERF** | Established Risk Factor? | Diagnosis / Condition |
| **G/E** | Gastroenteritis | Diagnosis / Condition |
| **LPR** | Laryngopharyngeal Reflux | Diagnosis / Condition |
| **PID** | Short for pelvic inflammatory disease (PID) is an infection of the female reproductive system, which includes the womb, fallopian tubes and ovaries. | Diagnosis / Condition |
| **RTI** | Respiratory Tract Infection | Diagnosis / Condition |
| **URTI\*** | For Upper Respiratory Tract Infection | Diagnosis / Condition |
| **AMOXYL\*** | Amoxicillin (recommended in MTG for treatment of pneumonia in children) | Drug |
| **ASA\*** | Aspirin | Drug |
| **CTX** | CTX usually stands for ceftriaxone, an antibiotic used to treat various bacterial infections. But in Malawi professionals suggested that this usually stands for co-trimoxazole commonly utilised in medical settings for the treatment of respiratory infections, urinary tract infections, and certain types of pneumonia, particularly in immunocompromised patients. | Drug |
| **DCN** | Doxycycline is an antibiotic that is commonly used to prevent malaria. | Drug |

| **Notation** | **Meaning of abbreviation** | **Type** |
| --- | --- | --- |
| **DETROLAM** | DETROL LA treats the symptoms of overactive bladder (leaks, strong sudden urges to go, going too often). | Drug |
| **DEXA** | Dexamethasone | Drug |
| **GENTA\*** | Gentamicin, a broad spectrum antibiotic used to treat severe bacterial infections, it is usually administered intravenously.<br>(recommended in MTG for treatment of pneumonia in children) | Drug |
| **METRO** | Stands for metronidazole is an antibiotic that is used to treat a wide variety of infections. | Drug |
| **NEO DEXA** | Neomycin / dexamethasone. Neomycin is an antibiotic. It is used to treat bacterial infections. Dexamethasone is a steroid. It is used to treat the inflammation associated with bacterial infections of the eye. | Drug |
| **PCM\*** | Short for “paracetamol” | Drug |
| **SP\*** | Drug used for malaria (sulphadoxine-pyrimethamine (SP) Date on the HP where this was used was 2006 | Drug |
| **FBC\*** | Full Blood Count, a type of blood test | Laboratory |
| **HCG\*** | Tumour markers -HCG (Can also be interpreted as pregnancy test) | Laboratory |
| **MRDT\*** | Malaria Rapid Diagnostic Test | Laboratory |
| **NEG** | Negative | Laboratory |
| **PITC\*** | HIV test, stands for Provider-Initiated HIV Testing and Counselling, E.g. PITC neg | Laboratory |
| **POS** | Positive | Laboratory |
| **VDRL\*** | "Venereal Disease Research Laboratory (VDRL)" test is a blood test designed to assess whether you have syphilis, a sexually transmitted infection (STI). E.g. VDRL neg | Laboratory |
| **BD\*** | Twice a day, for example, DCN 100mg bd | Pharmacy |
| **OD** | Once a Day, OD 0.2.5 - 4/6 | Pharmacy |
| **P/C\*** | p.c: after meals (Latin: post cibum). Indicates to take the medicine after a meal. | Pharmacy |
| **TDS\*** | Three times a day | Pharmacy |
| **TDT** | Three times daily | Pharmacy |
| **BF** | BF = Breathing Frequency | Vital Signs |
| **O/S** | Oxygen Saturation | Vital Signs |
| **PFT** | Pulmonary Function test, E.g. BF :+PFT SEEN 6050 | Vital Signs |
| **TEMP** | Temperature, e.g. TEMP: 37.8 | Vital Signs |
| **WGT** | Weight, e.g. WGT:6.8KG | Vital Signs |

Diagnoses were surprisingly few: sepsis, malaria, RTIs, and skin conditions appeared frequently. Some were written in full, such as sepsis, while others, particularly respiratory conditions, were often abbreviated, e.g. ARI for Acute Respiratory Infection. Reference to social history such as smoking habits, or to occupation were very few: one such note referred to voice loss suffered by a teacher. Family history and past medical history were not included in most visit notes.

The notes did not contain details of the physical examination of patients or history of illnesses. Reference to anatomy was made in a summary way with few descriptions. The was little to no reference to procedures such as CT scans or x-rays. This is consistent to the fact that most clinics do not have access to testing facilities. Most tests mentioned were rapid tests except for a few which may have been recorded in private clinics with functioning laboratories.

There was a higher uniformity in the recording of antibiotic drugs. Names of common drugs and their abbreviations appear to be well known to healthcare professionals: Amoxyl and CTX. Some drugs appear in their full name such as Artesunate; artesunate is the first-line treatment for children or adults with severe malaria.

#### Use of signature and stamps

We observed various ways in which practitioners signed their notes in HPs: some used only their signature, while others used their initials or a combination of their initials and surname. All were in lowercase letters. None used capital letters, a standard that is considered good practice in many medical settings. Some notes did not have any signature at all. However, our findings could be limited by the fact that we only captured a sample of pages sometimes one from each passport.

Private clinics and those run by the Christian Health Association of Malawi, a private organisation, used stamps. For example, at the H. Paker facility, stamps were used for verification to confirm that a patient has gone through data capturing before receiving treatment at the pharmacy, mainly on weekdays during daytime. The stamp was also used when referring patients to a central hospital, as part of the protocol for accessing tertiary care. The referral procedures requires that patients must first be seen by their local facility who can then refer them a central hospital.

#### There was a lack of guidelines and policies on medical notes recording

Participants did not receive any specific training on the use of HPs. One participant referred to consulting international books on good practice regarding communication with patients, but no national or specific guidelines for recording patient notes in HPs exist in Malawi. Some participants felt that the general note-taking skills learned in school should be sufficient. The Malawi Standards for Treatment Guidelines (MSTG) contains guidelines for prescribing medication that include a list of common medical abbreviations for drugs and conditions, and guidelines for in-patient prescriptions (19). In contrast to registers, where a defined list of abbreviations for medical conditions was consistently used, the diagnoses recorded in HPs were less standardized.

### Language Use

Entries in all medical records are done in English (9). The language for expressing symptoms, conditions and diagnosis are mastered in English and not in local languages though the conversation with patients is to be done mostly in local languages.

Participants held various opinions concerning the most convenient language to be used in making entries in health passports so that the content remains beneficial to both the patient and the health provider.

1. Making entries in Chichewa will not solve problem of communication because not all patients are fluent in Chichewa or know medical terms in Chichewa. It would also require that health professionals learn and or understand Chichewa for medical terms.
2. Many medical terms (in English language) do not have direct equivalent terms in Chichewa hence they translate to long descriptions that will require more space on paper; therefore, HPs with a small number of pages would fill up very quickly. Making entries in Chichewa will also be time more consuming hence will affect the number of patients to be treated in a day.
3. Differences in dialects: some local language term might have multiple meanings depending on the tribe, region or place. For instance, in Nkhotakhota among the Tonga tribe, ***ntchofu*** means hernia while in common Chichewa, ***ntchofu*** means stomach discomfort that results into nausea and vomiting yellow mucus.
4. Chichewa language may not precisely capture subtle differences between medical conditions as it can be done in English. For example, it will be difficult to express differences in different types of meningitis.
5. Multilingualism: Not all doctors will be able to know the language of the patient and the use of an interpreter may be needed. Preference for Chichewa may be construed as a political move to advance the language over other local languages.
6. The names of drugs will remain difficult to have them directly translated into Chichewa because they are *proper nouns* either in English, scientifically label or in the language of the country of manufacture.
7. There is no need to have the entries written in local language because health passports are more important to the health provider/ the doctor than to the patient. It is a communication tool among healthcare providers and not meant for the patient and that is why it is full of medical symbols than full medical expressions.

The doctor should take the responsibility of explaining the entries made in English in the language of the patient being treated. The diagnosis should appear in the local language alongside the English term.

> *“It will be good to translate the diagnosis written in the HP to a language that the patient understands.”*

> *“Patients will only just see the medicine given such as PCM Panadol, metronidazole etc. The majority do not know what the doctor’s conclusive diagnosis is as drawn from the symptoms. They don’t know what they are suffering from, so it will be good if entries are put in local languages because the health worker will be able to elaborate the diagnosis to the patient and the patient will be able to understand, in local terms, the diagnosis, that is written in his/her health passport.”*

Having access to interpreters was looked at favorably.

> *“A translator would ease the communication between health workers and clients”. (…) It will be more work for the health workers to learn the local languages, but this will benefit the client.”*

However, the presence of a translator or interpreter may inhibit patients:

> *“the moment you add a third party to the conversation, the patient may feel uncomfortable, a lot of Malawians value privacy.”*

The interpreter should be a health professional who is specifically trained to interpret. Patients should be asked to consent to an interpreter. Perhaps applications of AI and technology can offer a better solution to using human interpreters:

> *“A digital translator would be more preferable than a third party because that would be a good communication direct between clinicians and clients.”*

#### Participants did not support staff deployment based on language competence

Deploying medical personnel based solely on their language proficiency was not seen as an ideal solution to language barriers. Some health professionals preferred not to work in their home areas where their mother tongue was spoken, fearing a lack of respect from those who knew them. Additionally, such deployment was seen as unintentionally reinforcing tribalism, stereotypes about literacy levels, and cultural beliefs. Others worried that working in their home village or town may lead to increased absenteeism or lateness, as personal obligations could take precedence over their professional responsibilities.

### Impact of Literacy

Low literacy was an important factor that affected the quality of the information patients received. A cultural preference for verbal communication over written notes by healthcare professionals was acknowledged. Professionals should provide good explanations to patients and ensure they understand the consequence of their actions especially because some patients are afraid to ask follow-up questions. But many patients were not able to repeat the instructions given to them. Good rapport with the medical professional was very important. Most illiterate people had low self-esteem and feared when they meet clinicians hence they struggle to express themselves for these reasons. Literate patients were given better attention and were received more favourably by healthcare professionals than are illiterate people.

> *“There is a problem in the way health professionals communicate with those who come for medical help, at the clinics. Healthy workers provide half -baked information and that after physical examinations, observations, medication and diagnosis, we don’t ask for feedback from the patients. The illiterates remain the main victim of this because majority of them never understand what is written in their health books.”*

On the other hand, literate and educated people could become difficult like some who researched their conditions on the internet and were forceful in asking for specific medication from health professionals. Patients who attended private hospitals sometimes sought care for trivial reasons (“they are not really sick”) and were more interested to ask for specific medications they read about.

### Privacy

Both language and privacy were equally important to the quality of care. Poor building structures and planning often compromised patient privacy during consultations. Larger clinics had few consultation rooms, while smaller facilities lacked them entirely. In some cases, children under five were treated outdoors, standing in a queue. In some clinics, consultations took place in waiting areas, sometimes near antenatal services where children were being weighed. Privacy was further compromised when consultation room doors remained open during patient visits. In extreme cases, benches for waiting patients were positioned less than a meter from where consultations occurred, making it easy for those waiting to overhear sensitive medical discussions from the consultation room. Due to inadequate road infrastructure, clinics were often built near major roads. While this improved accessibility, it also created privacy concerns. For example, at Machinjiri Clinic, the maternity ward was so close to the street that passersby could hear women in labour.

## Discussion

In 2009, two doctors working and living in Malawi noted on the importance of HPs, saying that *“the health passport is treated with reverence*” by patients and that *“the booklet provides a complete and integrated record of immunizations, preventive health care priorities, major medical morbidities and a continuation record of clinical encounters”* (20). They expressed enthusiasm that patient-held records empower patients in managing their own healthcare and promote transparency in the practitioner-patient relationship, strengthening trust. However, as we have shown, today several factors hinder these benefits especially in rural areas and for poorer population. These included low patient literacy, language barriers, and inconsistent or unclear record-keeping by practitioners.

Most facilities in Zomba District, most especially rural government clinics, did not keep out-patient files on their premises, a situation that was also described by Tough in the districts of Chikwawa and Blantyre in 2018 (2). Due to gaps in medical history that is recorded in HPs, or inadequate use by health professionals, HPs are less able to support healthcare professionals in reaching a better or quicker diagnosis. HPs are also important when patients are referred to larger government hospitals in urban areas. The role of HPs as an educative and informative tool for patients is also uncertain because patients struggle to understand what is written in their notes due to low literacy and poor legibility and coding practices.

The good news is that the foundational role of HPs can be restored through simple and practical measures like those implemented to improve data collected in registers. Encouraging health professionals to use HPs more efficiently and educating patients on the importance of HPs may result in turning around the current situation. Removing barriers such as costs is also important for ensuring equitable access to health (21). Attention should be given also to addressing inequalities that may have been introduced unintentionally by programs that prioritise one segment of the population over other. This has led to some patients placing more value on HPs than others, for example women and under 5, rather than as men or young people.

There are no guidelines for keeping and writing patient records like those that exits in South Africa or Zimbabwe (22,23). Bad practices in record keeping are also a consequence of poor resources allocated for this task including training and supervision. Issues such as high workload and understaffing act as significant barriers that affect the quality of the records. Improving the availability of staff is not sufficient on its own, and regular training in records keeping, data use and analysis is needed (7). Health practitioners, regardless of whether they work in rural and under resources clinics, should have good access to information and comprehensive training that includes the correct use of medical terms, best practices for patient identification, and guidelines for using abbreviations and signatures.

There have been some initiatives to improve the design of HPs so that they collect better data, but these have been mainly for maternal and child booklets. The child health booklet was revisited in 2023 to improve the recording best practice for head circumference measurements necessary for the early detection of hydrocephalus (24). Other such efforts were found that the HPs used in Malawi lacked clear instructions for healthcare providers on standard care and condition management and were of limited informational value to pregnant women (25).

It must be recognised that, over the last years, health authorities in Malawi have neglected record keeping especially in rural areas. The most recent NHIS dates from 2015 (1) and the most recent National Health Policy was published in 2020 (26). Only NHIS has a dedicated section on the use of health passports despite their significance. However, HPs continue to be used in data verification for national statistics by checking and reconciling figures between HPs, registers and electronic systems (27). When discrepancies exist between HPs, EHRs, and facility registers, decision making is severely impacted, wrong numbers may lead to wrong resource allocation for programs related to neonatal health, malaria, immunization, and maternal care (28).

The shift towards eHealth and digital data capture may have led to a reduced attention to paper-based records such as HPs in policies and guidelines published after 2015. The NHIS states that *“Electronic Medical Records system shall be gradually introduced in all health facilities nationwide. When a fully functional EMR is introduced at a health facility, this shall be the primary data source.”* Perhaps the emphasis on digitalisation and less on digitising is misplaced given the established use of HPs and the advantages that they still bring to patients and professionals despite challenges. There is a need to address questions such as: What happens to the paper-based documents already collected in many facilities? What is worth digitising? How could lessons learnt from the use of paper based records inform the utilisation of digital systems? Will born-digital records overcome the challenges observed with paper-based HPs? These questions are widely relevant given that it is not only in developing countries where a switch from paper-based records to EHRs is under-way.

The use of health passports is not just about data recording but carries legal and ethical implications, such as records ownership and access to information, the use of medical records in court, and proper documentation and informed consent. In recent years, the right of patients to access their records has come under focus in many countries (29). In Malawi, such patient rights especially for the poor and in rural areas can be facilitated using HPs and thus improving their effectiveness should be a priority.

The written and the verbal communication are both important. Effective informed consent depends on clear communication with the patients, whether verbally or in writing. There is a need to invest in adequate resources for local language and communication for both patients and professionals. Addressing patient illiteracy and language barriers is essential to ensure that medical information is safeguarded, informed consent is possible and that patient records serve their intended purpose in healthcare. Participants cast doubts about the feasibility of writing notes fully in local languages such as Chichewa but saw value in writing at least the diagnosis in a language patients can understand. The use of interpreters was considered a possible solution to language barriers, but concerns were raised about privacy, potential loss of meaning during translation, and cultural resistance. These findings agree with studies from other African countries (30).

## Conclusion

Despite the rise of eHealth initiatives in Malawi, HPs and other paper-based records remain a vital tool in the healthcare system. HPs have retained their role as integrated, portable health records but their effectiveness is diminishing due to incomplete records in note taking and a low rate of utilisation of the patient history by health professionals. Despite these challenges, HPs provide the only record of care for the majority of people in Malawi. They occupy a well-established role in the process of medical consultation as a means of communication with the patient, but also with other professionals. While HPs are valued as patient-retained records, issues such as low patient literacy, language barriers, inconsistent and poor legibility, and a shift toward digital health systems have all limited their effectiveness. Time constraints and staff workload often prevent healthcare professionals from providing sufficient information on HPs.

It seems the government and ministries of health are overlooking the importance of HPs. There is an urgent need to improve record-keeping practices. Efforts should focus on restoring patient trust in the importance of health passports, developing language resources, standardising documentation, and utilising interpreters where necessary. Addressing these gaps will enhance patient understanding, continuity of care, and the overall effectiveness of HPs in Malawi’s healthcare system.

Although this research improves our understanding of the use and effectiveness of HPs in Malawi it does not cover all relevant dimensions in depth. We were also limited by time and resources in the geographical coverage of the study. Future work could investigate how decentralisation of health systems could potentially improve record keeping and data use at facility and district levels ensuring that HPs remain a valuable tool in both traditional and digital healthcare settings. As new sophisticated technologies are incorporated into health, rural clinics should not be left behind. HPs can provide value and can help ensure data quality, enhance patient care, and achieve effectiveness in decision making.

## Abbreviations

ART: An&retroviral therapy
EHRs: Electronic Health Records
FGD: Focus Group Discussion
HMIS: Health Management Informa&on Systems HPs Health Passports
LMIC: Lower and Middle Income Countries
MSTG: Malawi Standards for Treatments Guidelines
NHIS: Na&onal Health Informa&on System Policy
OPD: Out-pa&ents department

## Data Availability

All data produced in the present study are available upon reasonable request to the authors. The data is kept in a repository on the Open Science Framework ofs.io.

